# Exposure profiles of social-environmental neighborhood factors and psychotic-like experiences

**DOI:** 10.1101/2024.08.21.24312315

**Authors:** Benson Ku, Qingyue Yuan, Grace M. Christensen, Lina Dimitrov, Benjamin Risk, Anke Huels

**Affiliations:** Department of Psychiatry and Behavioral Sciences, Emory University School of Medicine, Atlanta, GA, USA; Department of Epidemiology, Rollins School of Public Health, Emory University, Atlanta, GA, USA; Department of Biostatistics and Bioinformatics, Rollins School of Public Health, Emory University, Atlanta, Georgia, USA; Gangarosa Department of Environmental Health, Rollins School of Public Health, Emory University, Atlanta, Georgia, USA

**Keywords:** Neighborhood characteristics, physical activities, psychotic-like experiences, social determinants of health, team sports

## Abstract

**Importance:** Recent research has demonstrated that domains of social determinants of health (SDOH) (e.g., air pollution and social context) are associated with psychosis. However, SDOHs have often been studied in isolation.

**Objective:** To identify distinct exposure profiles, estimate their associations with persistent distressing psychotic-like experiences (PLE), and evaluate whether involvement with physical activities partially explains this association.

**Design, Setting, and Participants:** This population-based study used data from the Adolescent Brain and Cognitive Development (ABCD) Study. Participants were recruited from 22 US sites between September 2016 and January 2022. Data from baseline and three follow-ups were included.

**Exposures:** Area-level geocoded variables spanning various domains of SDOH, including socioeconomic status (SES), education, crime, built environment, social context, and crime, were clustered using a self-organizing map method to identify exposure profiles.

**Main Outcomes and Measures:** Persistent distressing PLE was derived from the Prodromal Questionnaire-Brief Child Version across four years. Generalized linear mixed modeling tested the association between exposure profiles and persistent distressing PLE as well as physical activities (i.e., team and individual sports), adjusting for individual-level covariates including age, sex, race/ethnicity, highest level of parent education, family-relatedness, and study sites.

**Results:** Among 8,145 participants (baseline mean [SD] age, 9.92 [0.63] years; 3,868 (47.5%) females; 5,566 (68.3%) White, 956 (11.7%) Black, 159 (2.0%) Asian, and 1,480 (18.4%) Hispanic participants), five exposure profiles were identified. Compared to the reference Profile 1 (suburban affluent areas, 2521 children, 30.9%), Profile 3 (rural areas with low walkability and high ozone; 1459 children, 17.9%; adjusted OR: 1.34, 95% CI: 1.09—1.64) and Profile 4 (urban areas with high SES deprivation, high crime, and high pollution; 715 children, 8.8%; adjusted OR: 1.40, 95% CI: 1.08—1.81), were associated with persistent distressing PLE. Team sports mediated 6.14% of the association for Profile 3.

**Conclusion and Relevance:** This study found that neighborhoods characterized by rural areas with low walkability and urban areas with high socioeconomic deprivation, air pollutants, and crime were associated with persistent distressing PLE. Further research is needed to explore the pathways through which different environmental factors may impact the development of psychosis.

## Introduction

Psychotic-like experiences (PLEs), also referred to as subclinical psychotic symptoms or psychotic experiences, are odd or unreal perceptions, thoughts, or beliefs. ^1^ It is one of the earliest manifestations of psychotic disorders and can be common among children. ^2,3^ Although PLEs have been considered mild expressions of psychosis liability, PLEs that are persistent and distressing may be more indicative of psychopathology later on in life with greater functional impairment, greater cognitive impairment, and increased mental health service utilization. ^4,5^ Given the high prevalence of PLE in early adolescence and its clinical significance to later risk for psychosis and general psychopathology, exploring the risk factors and mechanisms underlying persistent and distressful PLE is crucial to improve psychiatric risk assessments and more effective prevention strategies in children and adolescents.

It has been long known that urban upbringing is a major risk factor for developing psychotic disorders, including schizophrenia. ^6–8^ Under the social determinants of health model (SDOH) for psychosis, certain environmental and social conditions (e.g., air pollutants, social fragmentation, and socioeconomic deprivation) may impact downstream psychosocial and biological processes and, in turn, lead to psychosis. ^9,10^ Early exposure to air pollution has been shown to be associated with increased odds of psychotic-like experiences in adolescence, potentially through inflammation and excessive oxidative stress. ^11–13^ The Social Disorganization Theory posits that the wider community plays an important role in shaping social interactions, whether through sports or extracurricular activities, which may be crucial for mental health, especially for children. ^14,15^ For example, youth living in neighborhoods that were more walkable, had less crime, and greater social cohesion participated in more physical activities, particularly team sports. ^16–18^ And greater involvement with team sports, as opposed to individual sports, was shown to be associated with reduced risk for future psychopathology, including persistent distressing PLE. ^19–21^ Perhaps structural factors may facilitate youth coming together to play team sports, and certain aspects of these social ties may be relevant to offset the risk of psychosis. ^21^

Prior literature has largely focused on analyzing each neighborhood characteristic as it relates to psychosis separately. ^22–24^ Existing measurement tools, such as the Area Deprivation Index (ADI) and Child Opportunity Index (COI), have been limited in their ability to capture the multifaceted impacts of various neighborhood characteristics simultaneously while also understanding individual contributions of each neighborhood characteristic. These indices are constructed from an arbitrarily chosen set of SDOH variables, and little is currently known about the combinations of these environmental and social factors as well as the mechanisms through which they lead to psychosis. Without such information, it may be difficult to devise effective, targeted policies and interventions.

This study aimed to fill this knowledge gap by using innovative methodologies that capture and analyze SDOH’s multidimensional nature and uncover its associations with distressing PLE and physical activities. We clustered area-level exposures based on various indices of air pollutants and psychosocial factors into exposure profiles. Then we investigated whether these exposure profiles would be associated with persistent distressing. We also tested team sports as a potential mediator of these associations. We hypothesized that certain urban exposure profiles would be associated with persistent distressing PLE and that team sports would partially explain this association.

## Methods

The data were collected from a population-based sample of 9-to-10-year-olds in the Adolescent Brain and Cognitive Development Study (ABCD) 5.0 release, which included visits collected between September 1, 2016, and January 15, 2022. ^25^ Data from baseline and three follow-up time points were included.

The ABCD Study is a nationwide longitudinal study of brain, behavioral, and cognitive development in adolescents and involves 22 research sites throughout the United States, with more than 11,868 children recruited at baseline. Within these sites, public, private, and public charter schools within a 50-mile radius of the data-collecting site were randomly selected for recruitment. ^26^ General inclusion and exclusion criteria for the ABCD study have been described elsewhere. ^26–28^ Informed written consent was obtained from children and their parents, and ethical approval was obtained from each site’s institutional review boards. We followed the Strengthening the Reporting of Observational Studies in Epidemiology (STROBE) reporting guidelines.

### Participants

This study included 8,145 participants based on the availability of environmental exposures, outcome variables, and sociodemographic covariates. **eFigure 1** shows a flowchart of included and excluded participants. **eTable 1** compares those included and excluded based on missing data.

**Figure 1.**
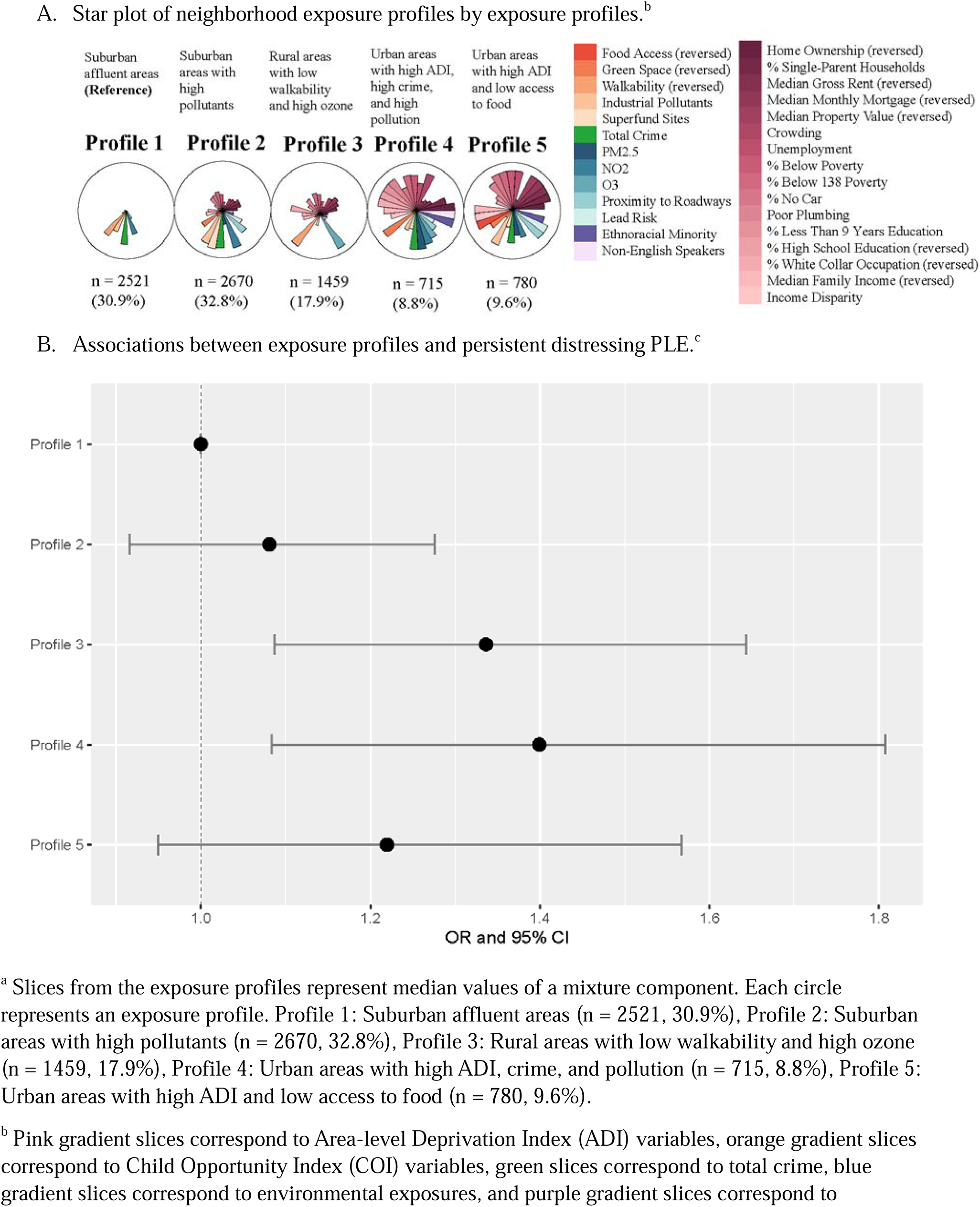
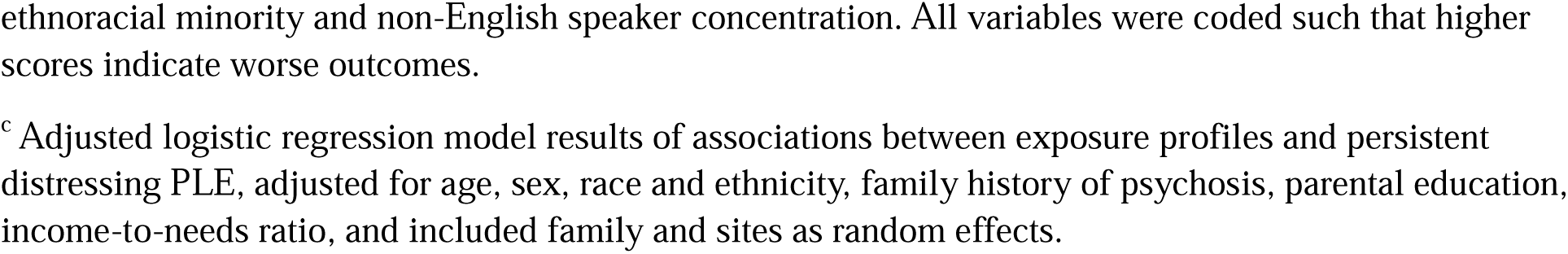
Associations between exposure profiles and persistent distressing PLE.^a^.

### Sociodemographic and clinical characteristics

Sociodemographic characteristics, including age, sex, race, ethnicity, parental education, household income, and family history of psychosis, were collected through parent reports and interviews during baseline assessment. Race and ethnicity were aggregated into 5 categories: non-Hispanic White, non-Hispanic Black, non-Hispanic Asian, non-Hispanic other races, and Hispanic. High parental education was defined as having at least one parent or caregiver who obtained a bachelor’s degree or higher. The income-to-needs ratio was calculated by the median value of the income band divided by the federal poverty line for the respective household size. ^29^ A value greater or less than one would denote above or below the poverty threshold, respectively. Family history of psychosis in first-degree and second-degree relatives was assessed using the parent-rated Family History Assessment Module Screener. ^30^

### Area-level exposures

Area-level data were derived from participants’ primary home addresses at baseline, which were geocoded to retrieve information at the census tract level. These geospatial location data were then linked to external environmental constructs, which were part of the ABCD 5.0 release. ^25^ Exposure variables were used to construct five domains, including the Area Deprivation Index (ADI), ^31,32^ Child Opportunity Index 2.0 (COI), ^33^ Crime, ^34^ Environmental Quality, ^35,36^ and Social Vulnerability Index (SVI), ^37^ based on their respective sources and prior literature. ^25^ A comprehensive list of variables included in our study, exposure definition, and years measured are described in **Table 1** and **eTable 2**. Distance to major roadways, population density, and percentage of households without a car, were used to classify the exposure profiles as urban, suburban, and rural as done in prior literature. ^38,39^

**Table 1.**
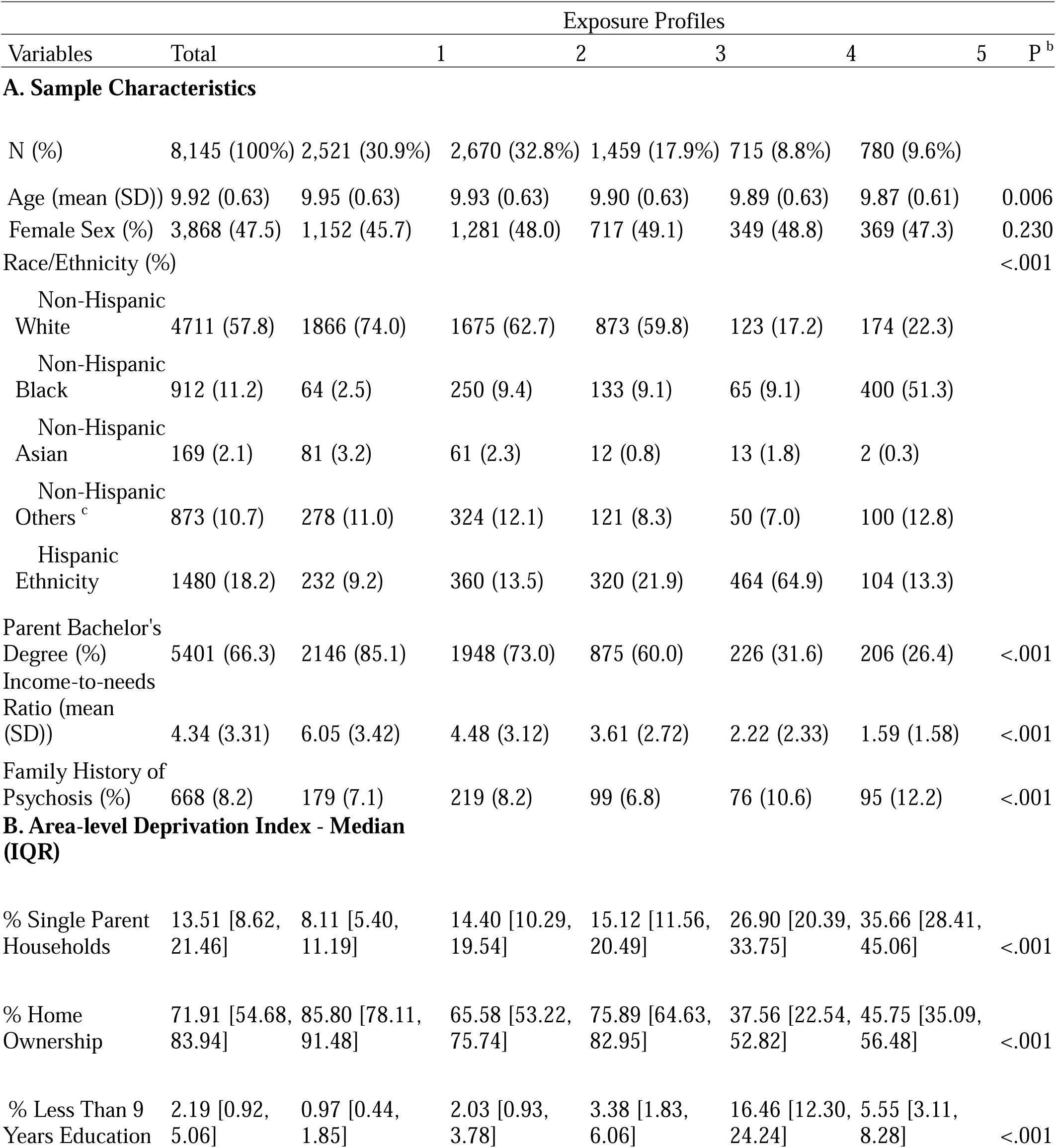
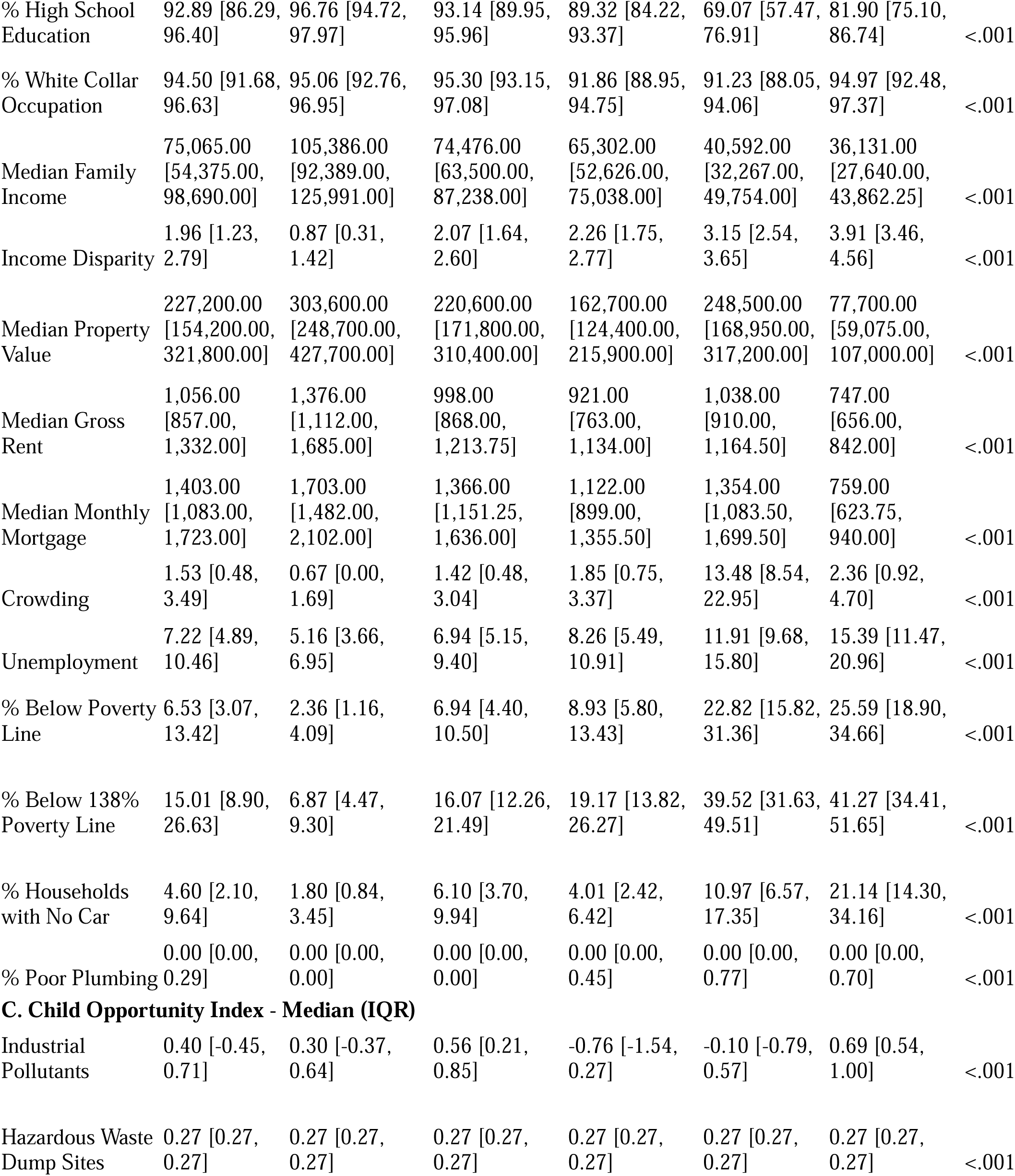
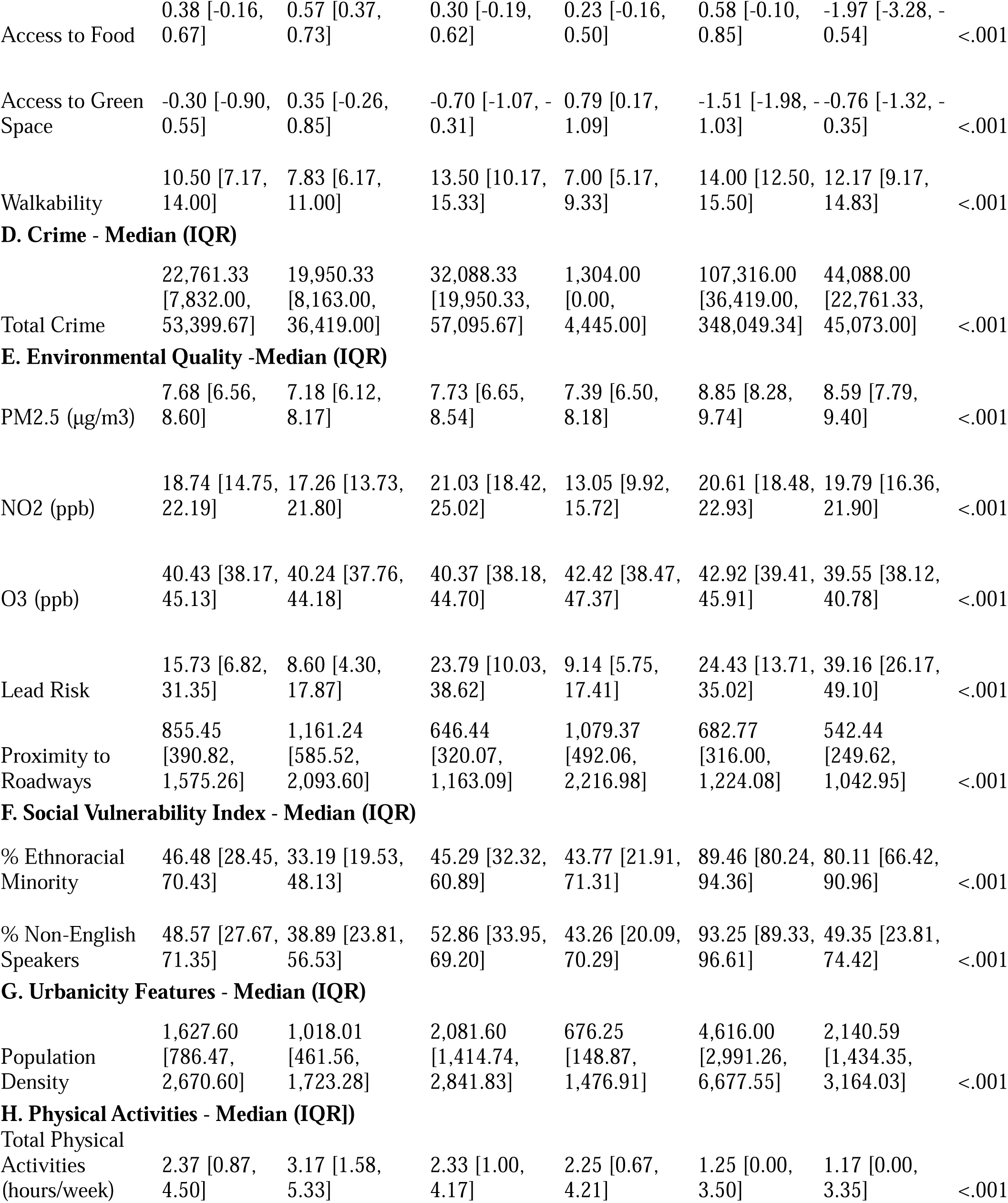
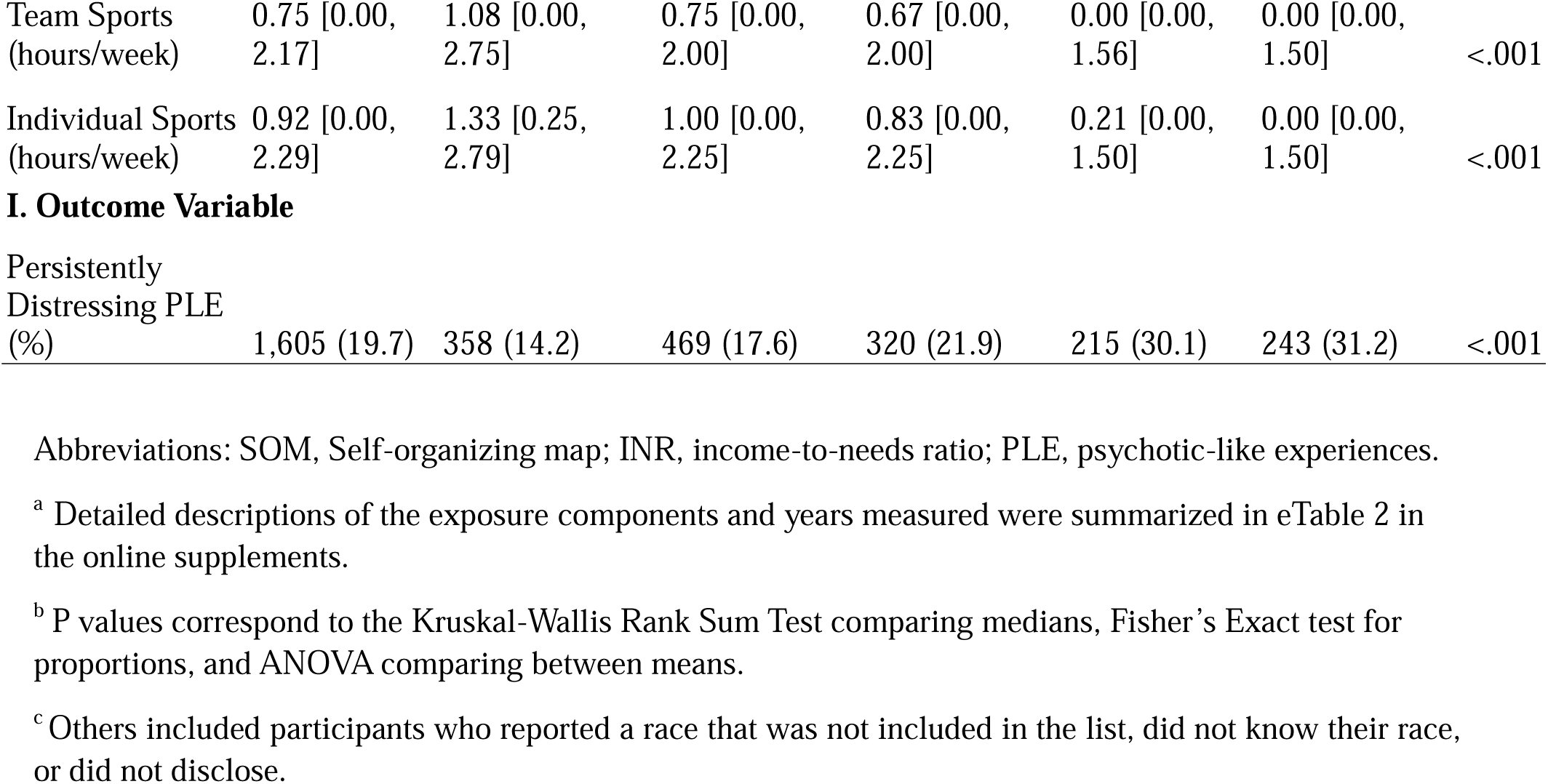
Description of Study Sample by Exposure Profiles. A. Study Sample Characteristics. B. Area-level Deprivation Index. C. Child Opportunity Index. D. Crime. E. Environmental Quality. F. Social Vulnerability Index. G. Urbanicity Features. H. Physical Activities. I. Outcome Variable. ^a^

### Persistent distressing psychotic-like experiences (PLE)

The Prodromal Questionnaire-Brief Child Version (PQBC) was completed by participants to assess psychotic-like experiences (PLE) over the past month. ^40^ Initially, participants were categorized as either having experienced distressing PLE (rating of at least one PLE>= 3 on a five-point distress scale) or not having experienced distress, following criteria based on prior literature on distressing PLE. ^4^ Participants with distressing PLEs were grouped into persistent and non-persistent categories, with “persistent” indicating distress at two or more follow-ups out of 4 waves (baseline, 1-year, 2-year, and 3-year). This binary categorization was based on prior literature demonstrating such persistency (as opposed to transiency) was specifically associated with a polygenic risk score for schizophrenia, ^21^ greater cognitive deficits, worse social functioning, and more health service utilization^4^ as well as a greater risk for future diagnosis of schizophrenia. ^41^ Because psychotic symptoms may develop throughout adolescence, we conducted a sensitivity analysis and redefined persistent distressing PLE to incorporate individuals who reported distressing PLE only at the last follow-up.

### Physical activities

Physical activities were derived from the Sports and Activities Involvement Questionnaire (SAIQ). Physical activities were further subcategorized into team and individual sports (**eTable 3**). The study focused on the level of involvement in 23 physical activities, with caregivers reporting (1) the time spent per session in minutes, (2) the number of days per week of participation, and (3) the number of months per year of engagement. The mean participation hours per week for each sport endorsed in the past 12 months were calculated using the formula: average hours per week per sport in the past year = (time spent × days per week × months per year) / 12 / 60. ^42^

### Statistical analysis

The self-organizing map (SOM) is an unsupervised machine learning technique for multi-dimensional data reduction and visualization, and identifying profiles of correlated characteristics (e.g., air pollutants and socioeconomic deprivation). SOM aims to map complex, multi-dimensional data into a simpler, easy-to-visualize format where multiple exposures are grouped into an exposure profile. SOM was used to cluster 29 area-level exposure profiles in this study. See **Table 1** and **eTable 2** for a complete list of exposure variables. The optimal number of profiles was determined using within-cluster sum of squares, between-cluster sum of square statistics, and visual inspection of the exposure profile star plot. ^43^ These methods seek to maximize homogeneity within profiles and heterogeneity between profiles. The reference profile had the highest socioeconomic status and lowest hazardous environmental exposure (e.g., air pollution). Once participants were assigned to an exposure profile, unadjusted and adjusted logistic mixed models were used to estimate the association between exposure profiles, modeled as a categorical exposure, and persistent distressing PLE as well as physical activities (i.e., team and individual sports). The model adjusted for age, sex, family history of psychosis, race and ethnicity, parental education, income-to-needs ratio, as individual-level fixed effects, and family groups and recruiting sites as random effects.

We then assessed whether team and individual sports would mediate the association between exposure profile and persistent distressing PLE. Mediation analysis was only conducted for exposure profiles that were statistically significantly associated with persistent distressing PLE by modeling each significant exposure profile as a dichotomous exposure variable in the mediation analysis. Detailed definitions and methodology for the mediation analyses were provided in the **eMethods**.

Subsequently, we used another method, weighted quantile sum (WQS) regression, to understand the overall effect of the exposure mixture and how much of the total mixture effect on the outcome is explained by each area-level characteristic within the mixture. WQS is a method that estimates the effect on the outcome of increasing all exposures simultaneously by one quantile, as well as the weighted contribution of each exposure variable to the association between the overall exposure mixture and the outcome while accounting for the complex correlation structure among exposures. ^44–46^ All SOM analyses were performed using code within the ECM package (https://github.com/johnlpearce/ECM). The other analyses used the following R packages, lme4, ^47^ lmerTest, ^48^ mediation, ^49^ gWQS. ^50^ All analyses were conducted using R version 4.2.1. Statistical significance was determined using an alpha-level of 0.05.

## Results

### Descriptive statistics

The study included 8,145 participants aged 9 to 10 years at baseline followed until 13 to 14 years, with 3,868 (47.5%) females, 5,566 (68.3%) non-Hispanic White, 956 (11.7%) non-Hispanic Black, 159 (2.0%) non-Hispanic Asian, and 1,480 (18.4%) Hispanic participants. Among the participants, 5,401 (66.3%) had parents with at least a bachelor’s degree and 668 (8.2%) had a family history of psychosis. Across the four years, 1605 (19.7%) participants had persistent distressing PLE (**Table 1**). All individual neighborhood-level characteristics correlate with each other in the expected direction (**eFigure 2**).

### Five distinct exposure profiles

Five exposure profiles were identified upon visual inspection. Each participant was assigned to one of the five profiles, and their demographics stratified by the exposure profiles are shown in **Table 1**. **Figure 1A** presents a detailed description of each profile.

### Associations between exposure profiles and persistent distressing PLE

Profile 3, characterized by rural areas with low walkability and high ozone, and Profile 4, characterized by urban areas with high ADI, crime, and pollution, were significantly associated with persistent distressing PLE compared to the reference Profile 1 (Profile 3: adjusted odds ratio (OR): 1.34, 95% CI: 1.09—1.64, p=.006; Profile 4: adjusted OR: 1.40, 95% CI: 1.08—1.81, p=.01) (**Table 2**, **Figure 1B**). Sensitivity analyses of (1) an alternative version of persistent distressing PLE and (2) only including participants who lived in their addresses for more than one year showed consistent results (**eTable 4**). Variance inflation factors (VIF) for exposure profiles and individual-level covariates ruled out multicollinearity in our models (**eTable 5**).

**Table 2.**
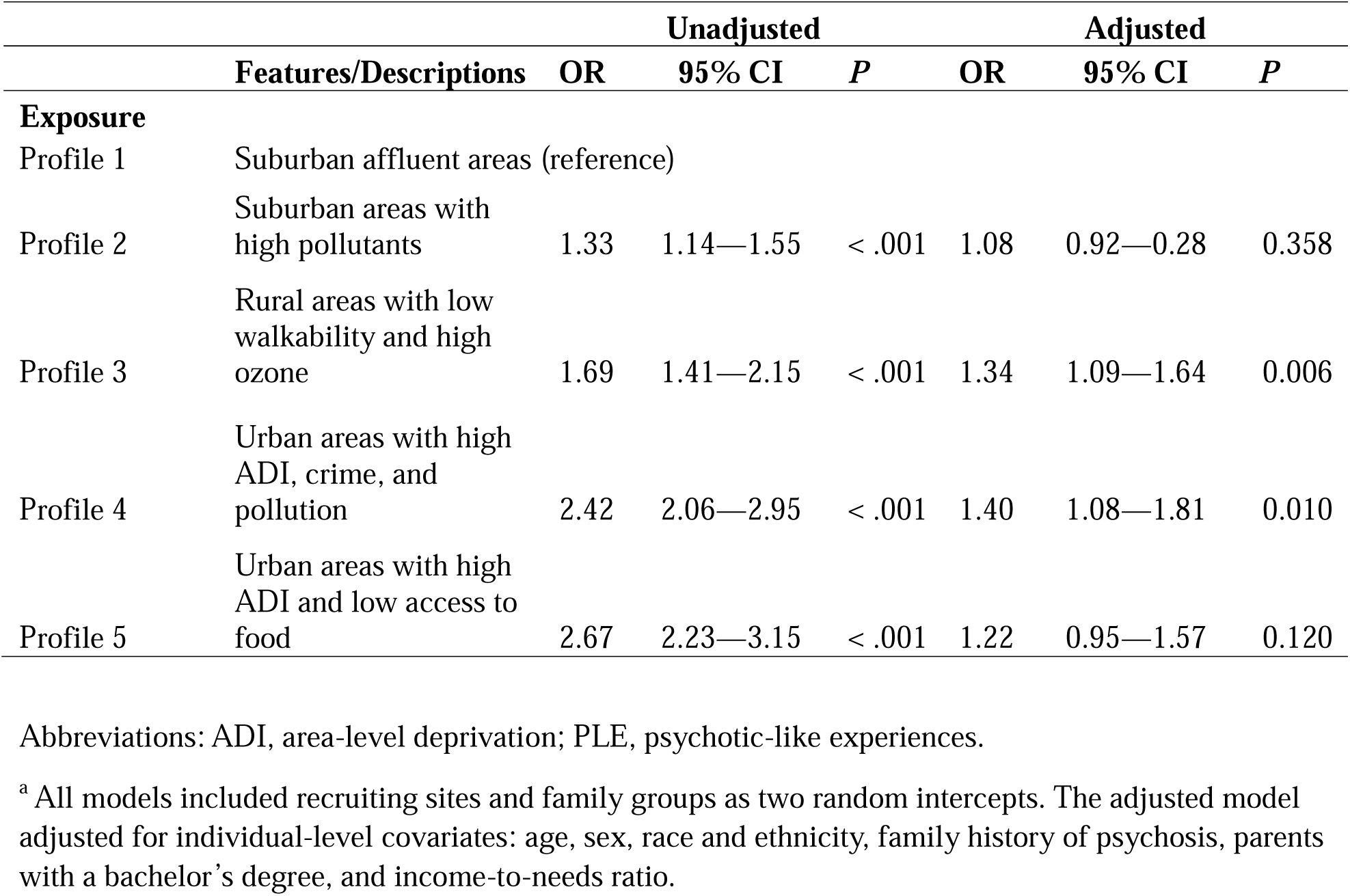
Associations between exposure profiles and persistent distressing PLE.^a^.

### Mediation of the relationship between Profile 3 and persistent distressing PLE by team sports

Team sports explained 6.14% of the relationship between Profile 3 and persistent distressing PLE (indirect effects adjusted β: 0.002, 95% CI: <.001—.005, p =.013) (**Figure 2**). However, team sports did not significantly mediate the association between Profile 4 and persistent distressing PLE (**Figure 2**). Total physical activities or individual sports did not significantly mediate the association between Profile 3 or Profile 4 and persistent distressing PLE (**eTables 6 and 7**).

**Figure 2.**
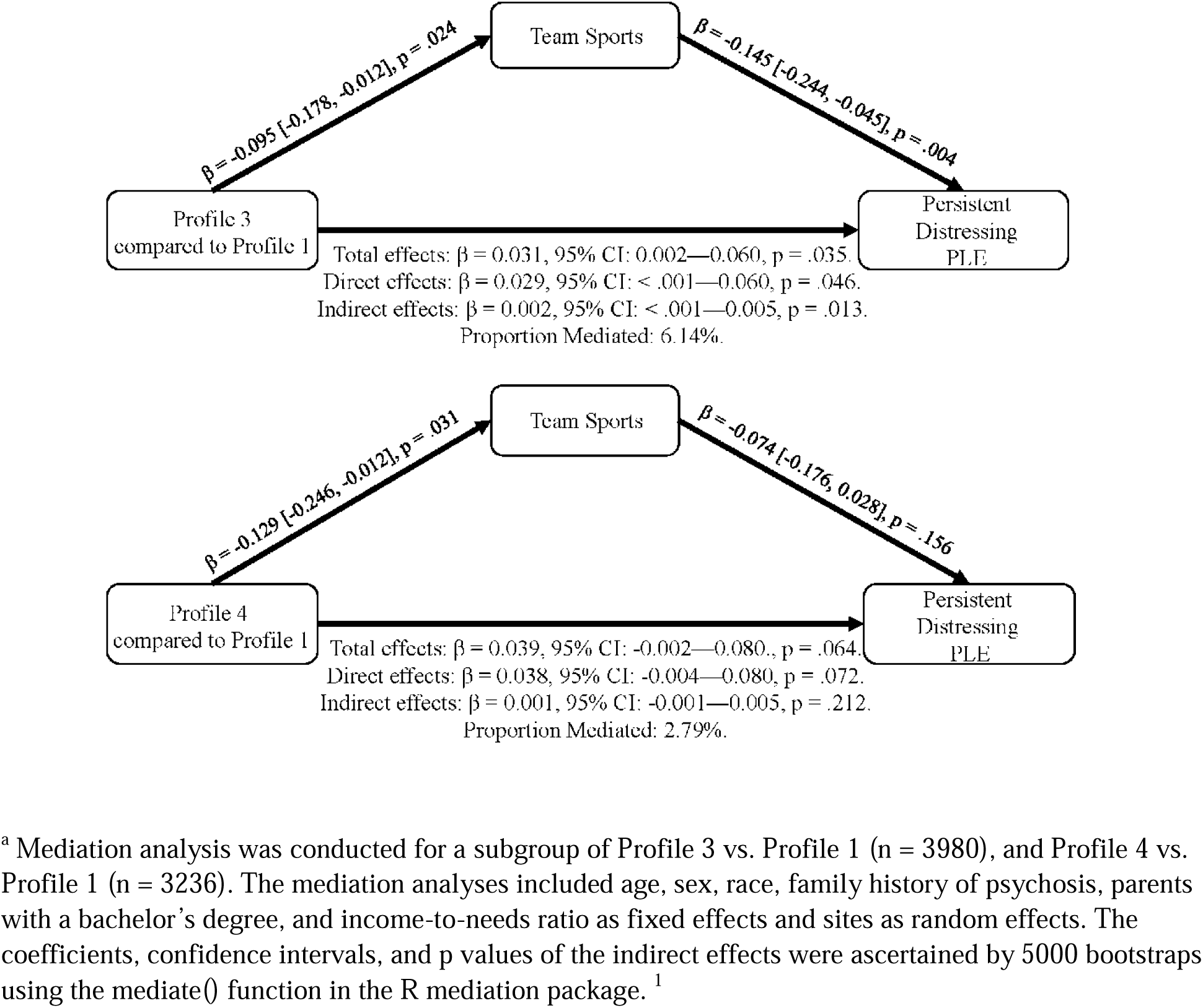
Mediation of the relationship between Profile 3 and persistent distressing PLE by team sports.^a^.

### Individual exposure contribution assessed by WQS regression weights

The positively constrained model demonstrated that the overall effect of increasing all exposures simultaneously by one decile corresponded to 12% greater odds for endorsement of persistent distressing PLE (adjusted OR: 1.12, 95% CI: 1.02—1.23, p=.03). The tree plot presented in **Figure 3** demonstrates the contribution of each exposure to a higher odds of persistent distressing PLE with greater total crime having the highest mean weight at 0.130, followed by lower walkability at 0.106, greater ethnoracial minority concentration at 0.090, higher ozone at 0.085, and greater industrial pollutants at 0.077 (**eTable 8**). The negatively constrained model was not significant (adjusted estimate: < .01, 95% CI: -0.10—0.09, p=.95) (**eTable 9**).

**Figure 3.**
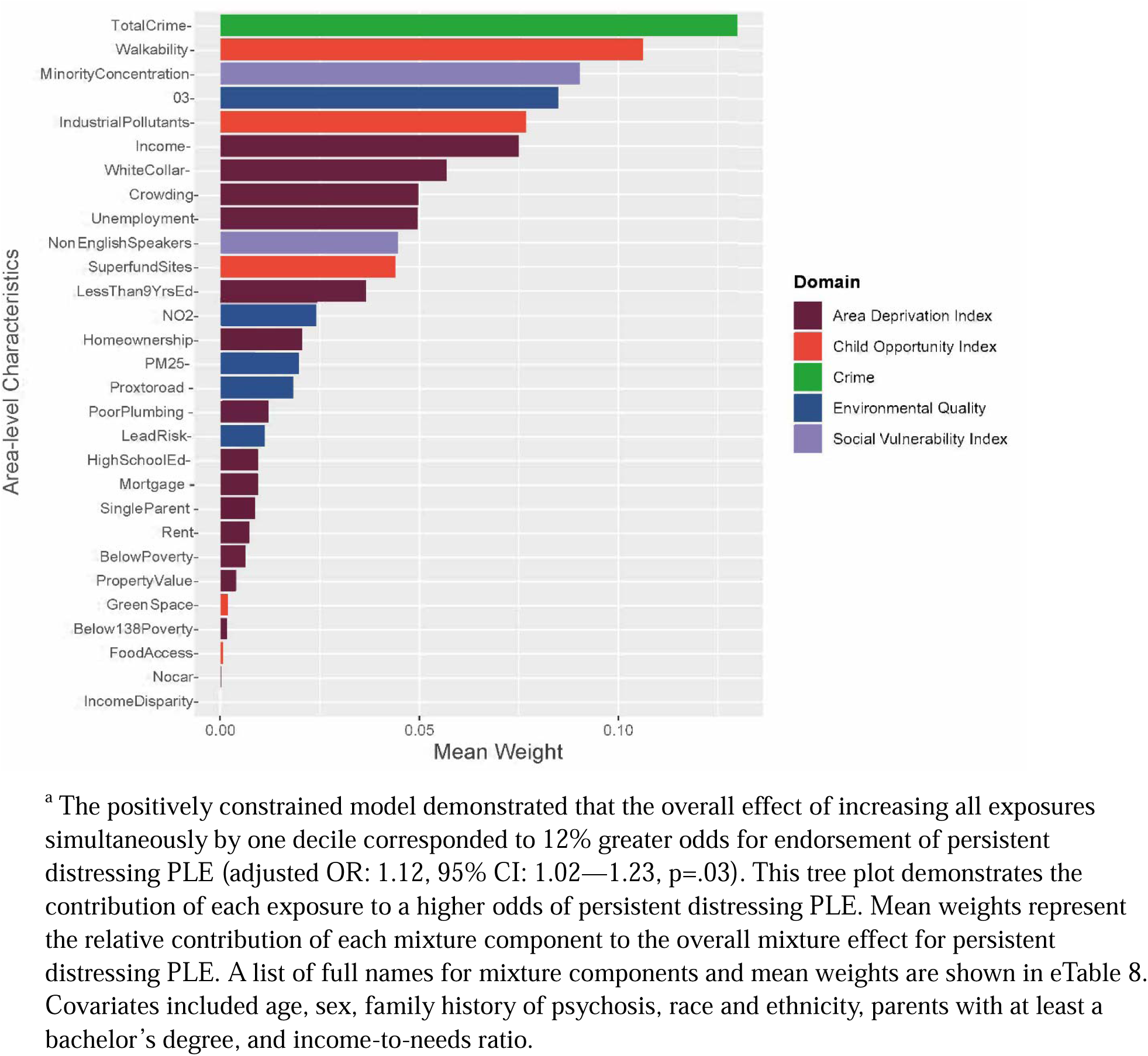
Positive Constrained Weighted Quantile Sum (WQS) Regression Weights. ^a^.

## Discussion

In this study, we identified five unique exposure profiles and investigated their associations with persistent distressing PLE and physical activities. We found two exposure profiles to be associated with greater odds of developing persistent distressing PLE. Rural areas that were less walkable with high ozone (Profile 3) and urban areas with high socioeconomic disadvantage, crime, and pollution (Profile 4) were both uniquely associated with greater odds for persistent distressing PLE even after adjusting for individual-level covariates. It is likely that a combination of factors, including financial stress, food insecurity, limited opportunities for physical activities, and the presence of air pollutants, might heighten the likelihood of PLE. ^13,51,52^ Chronic stress has been shown to disrupt the hypothalamic-pituitary-adrenal axis and elevate cortisol levels, which has been shown to be associated with neuroanatomical changes linked to psychotic illnesses. ^53–55^ Subsequently, our mediation analyses suggested that there may be distinct biopsychosocial pathways through which the environment may play a role in the development of psychosis. For example, the degree of involvement in team sports (but not individual sports) modestly mediated the association between Profile 3 and PLE by more than 6%. However, none of the subcomponents of physical activities mediate such relationships for Profile 4. Although the association between Profile 3 and PLE was mainly driven by other factors rather than team sports, it is possible that involvement in team sports (e.g., pick-up basketball) and perhaps other informal social interactions may be less likely to occur in more rural and less walkable places. ^56^ This lack of social engagement with peers may play a role in the future risk of psychotic experiences. ^57,58^

Our findings demonstrating that the association between Profile 4 and PLE was not explained by physical activity suggest that there may be an alternative mechanism through which urban areas characterized by high crime and pollution may be associated with PLE. It is possible that threat from exposure to crime, ^59^ air pollution, ^60^ and/or financial stress^61^ may play a more important role for this subgroup of youth with these specific combinations of exposures. In fact, recent literature has suggested that deprivation and threat may be two environmental factors that underlie partially distinct biological pathways in the development of psychosis. ^62,63^ Our findings may partially reflect this dimensional model of adversity in psychosis, and it is possible that different environmental factors may impact psychosis through distinct mechanisms.

In addition to the findings using SOM, the WQS regression model also points to similar factors that may drive the development of PLEs. High crime rates and low walkability carried the greatest weight in driving the effects of persistent distressing PLE, which align with high crime rates in Profile 4 and low walkability in Profile 3. Prior research has shown that living in high-crime neighborhoods was associated with subclinical psychotic symptoms, including suspiciousness and paranoia among help-seeking adolescents, ^64,65^ as well as a higher incidence of first-onset schizophrenia. ^59^ Although walkability has not been widely studied in psychosis, places with less walkability tend to have less access to community services and recreational centers. ^16,66,67^ Access to these resources may have downstream effects on exercise and involvement with team sports, which has been shown to be inversely associated with psychopathology and PLE. ^19,20,68,69^

One of the key strengths of this study is our approach to identifying multidimensional exposure profiles as measured by area-level characteristics, as opposed to traditional single-indexing methods. In addition, we used another exposure mixture method (i.e., WQS) to enhance the rigor of our findings and demonstrated consistent factors that may be driving the association with PLE. Future prospective studies should further investigate the biopsychosocial mechanisms through which neighborhood characteristics, including crime and walkability (etc. types of crime, park access, street connectivity, mixed land use) ^70^ may drive the development of psychotic disorders.

### Limitation

This study has several limitations. First, we excluded several participants due to missing data. Participants excluded were from lower SES households, which may impact the findings’ generalizability. Second, PLEs were self-reported, and physical activities were parent-reported, which may be biased. Third, this study did not test whether there was a sensitive period in which environmental factors may have a more pronounced effect on long-term psychosis risk. Prospective studies should collect this data and analyze whether environmental factors at various developmental periods may differentially impact psychosis risk.

### Conclusion

This study identified five exposure profiles, of which two were associated with persistent distressing psychotic-like experiences among children and adolescents across four years. These two exposure profiles were characterized by (1) rural areas with low walkability with high ozone (Profile 3), and (2) urban areas with high socioeconomic deprivation and high pollutants (Profile 4). Moreover, we found that less involvement with team sports partially mediated the positive association between Profile 3 (but not Profile 4) and greater odds of developing persistent distressing psychotic-like experiences. These findings suggest that there may be distinct mechanisms through which environmental factors may impact psychosis. Future studies should further explore the mechanisms through which living in certain neighborhoods may confer risk for the development of psychosis.

## Data Availability

All data from the Adolescent Brain Cognitive Development (ABCD) Study (https://nda.nih.gov/abcd/request-access) are made available to researchers from universities and other institutions with research inquiries following institutional review board and National Institute of Mental Health Data Archive approval.

## Key Points

### Question

What are the underlying complex patterns of the environmental and social determinants of health, and what are their associations with persistent distressing psychotic-like experiences among children in the US?

### Findings

Two distinct exposure profiles characterized by rural areas with low walkability and urban areas with high socioeconomic deprivation, air pollutants, and crime were associated with persistent distressing psychotic-like experiences.

### Meaning

Several adverse environmental and social factors were jointly associated with psychotic-like experiences. Further research is needed to investigate the mechanisms through which these factors impact the development of psychosis.

## Acknowledgments

This work was supported by the National Institute of Mental Health (NIMH K23-MH129684; Dr. Ku), the National Institute on Aging (NIA R01AG079170; Huels/Wingo), the National Institute of Mental Health (NIMH R01 MH129855; Risk), the Emory Constructive Collision Grant (Ku/Risk/Huels), and Emory HERCULES Pilot Award (Ku/Risk/Huels). The content is solely the responsibility of the authors and does not necessarily represent the official views of the National Institutes of Health, the National Institute of Mental Health, or Emory University.

## Data Sharing Statement

The ABCD study anonymized data are released annually and are publicly available via the NIMH Data Archive (NHA). All data from the Adolescent Brain Cognitive Development (ABCD) Study (https://nda.nih.gov/abcd/request-access) are made available to researchers from universities and other institutions with research inquiries following institutional review board and National Institute of Mental Health Data Archive approval.

## References

1. Hinterbuchinger B, Mossaheb N. Psychotic-Like Experiences: A Challenge in Definition and Assessment. Frontiers in Psychiatry. 2021;12doi:10.3389/fpsyt.2021.582392

2. Linscott RJ, van Os J. An updated and conservative systematic review and meta-analysis of epidemiological evidence on psychotic experiences in children and adults: on the pathway from proneness to persistence to dimensional expression across mental disorders. Psychol Med. Jun 2013;43(6):1133–49. doi:10.1017/s0033291712001626

3. Schultze-Lutter F, Renner F, Paruch J, Julkowski D, Klosterkötter J, Ruhrmann S. Self-reported psychotic-like experiences are a poor estimate of clinician-rated attenuated and frank delusions and hallucinations. Psychopathology. 2014;47(3):194–201. doi:10.1159/000355554

4. Karcher NR, Loewy RL, Savill M, et al. Persistent and distressing psychotic-like experiences using adolescent brain cognitive development study data. Mol Psychiatry. Mar 2022;27(3):1490–1501. doi:10.1038/s41380-021-01373-x

5. Oh H, Karcher NR, Soffer[Dudek N, Koyanagi A, Besecker M, Devylder JE. Distress related to psychotic experiences: Enhancing the world health organization composite international diagnostic interview psychosis screen. International Journal of Methods in Psychiatric Research. 2024;33(1)doi:10.1002/mpr.1977

6. Pignon B, Szoke A, Ku B, Melchior M, Schurhoff F. Urbanicity and psychotic disorders: Facts and hypotheses. Dialogues Clinical Neuroscience. Dec 2023;25(1):122-138. doi:10.1080/19585969.2023.2272824

7. Fan CC, McGrath JJ, Appadurai V, et al. Spatial fine-mapping for gene-by-environment effects identifies risk hot spots for schizophrenia. Nature Communications. 2018;9(1)doi:10.1038/s41467-018-07708-7

8. March D, Hatch SL, Morgan C, et al. Psychosis and place. Epidemiologic Reviews. 2008;30:84-100. doi:10.1093/epirev/mxn006

9. Karcher NR, Schiffman J, Barch DM. Environmental Risk Factors and Psychotic-like Experiences in Children Aged 9-10. J Am Acad Child Adolesc Psychiatry. Apr 2021;60(4):490–500. doi:10.1016/j.jaac.2020.07.003

10. Anglin DM, Ereshefsky S, Klaunig MJ, et al. From Womb to Neighborhood: A Racial Analysis of Social Determinants of Psychosis in the United States. Am J Psychiatry. Jul 2021;178(7):599–610. doi:10.1176/appi.ajp.2020.20071091

11. Newbury JB, Arseneault L, Beevers S, et al. Association of Air Pollution Exposure With Psychotic Experiences During Adolescence. JAMA Psychiatry. 2019;76(6):614. doi:10.1001/jamapsychiatry.2019.0056

12. Newbury JB, Stewart R, Fisher HL, et al. Association between air pollution exposure and mental health service use among individuals with first presentations of psychotic and mood disorders: retrospective cohort study. Br J Psychiatry. Dec 2021;219(6):678–685. doi:10.1192/bjp.2021.119

13. Newbury JB, Heron J, Kirkbride JB, et al. Air and Noise Pollution Exposure in Early Life and Mental Health From Adolescence to Young Adulthood. JAMA Network Open. 2024;7(5):e2412169–e2412169. doi:10.1001/jamanetworkopen.2024.12169

14. Ku BS, Compton MT, Walker EF, Druss BG. Social Fragmentation and Schizophrenia: A Systematic Review. J Clin Psychiatry. Dec 7 2021;83(1)doi:10.4088/JCP.21r13941

15. Sampson RJ, Groves WB. Community structure and crime: Testing social-disorganization theory. American journal of sociology. 1989;94(4):774–802.

16. Wang ML, Narcisse MR, McElfish PA. Higher walkability associated with increased physical activity and reduced obesity among United States adults. Obesity (Silver Spring). Feb 2023;31(2):553–564. doi:10.1002/oby.23634

17. Xiao Y, Mann JJ, Chow JC-C, et al. Patterns of Social Determinants of Health and Child Mental Health, Cognition, and Physical Health. JAMA Pediatrics. 2023;177(12):1294–1305. doi:10.1001/jamapediatrics.2023.4218

18. Wang ML, Narcisse MR, Alatorre S, Kozak AT, McElfish PA. Neighborhood social cohesion and physical activity and obesity outcomes among Native Hawaiian and Pacific Islander individuals. Obesity. 2022;30(1):249–256.

19. Noordsy DL, Burgess JD, Hardy KV, Yudofsky LM, Ballon JS. Therapeutic Potential of Physical Exercise in Early Psychosis. American Journal of Psychiatry. 2018;175(3):209–214. doi:10.1176/appi.ajp.2017.17060716

20. Hoffmann MD, Barnes JD, Tremblay MS, Guerrero MD. Associations between organized sport participation and mental health difficulties: Data from over 11,000 US children and adolescents. PLoS One. 2022;17(6):e0268583. doi:10.1371/journal.pone.0268583

21. Ku BS, Yuan Q, Arias-Magnasco A, et al. Associations Between Genetic Risk, Physical Activities, and Distressing Psychotic-like Experiences. Schizophr Bull. Aug 22 2024;doi:10.1093/schbul/sbae141

22. Erzin G, Gülöksüz S. The exposome paradigm to understand the environmental origins of mental disorders. Alpha Psychiatry. 2021;22(4):171.

23. Ku BS, Ren J, Compton MT, Druss BG, Guo S, Walker EF. The association between neighborhood-level social fragmentation and distressing psychotic-like experiences in early adolescence: the moderating role of close friends. Psychol Med. Feb 16 2024:1–9. doi:10.1017/S0033291724000278

24. Anglin DM, Espinosa A, Addington J, et al. Association of Childhood Area-Level Ethnic Density and Psychosis Risk Among Ethnoracial Minoritized Individuals in the US. JAMA Psychiatry. Dec 1 2023;80(12):1226–1234. doi:10.1001/jamapsychiatry.2023.2841

25. Fan CC, Marshall A, Smolker H, et al. Adolescent Brain Cognitive Development (ABCD) study Linked External Data (LED): Protocol and practices for geocoding and assignment of environmental data. Developmental cognitive neuroscience. Dec 2021;52:101030. doi:10.1016/j.dcn.2021.101030

26. Garavan H, Bartsch H, Conway K, et al. Recruiting the ABCD sample: Design considerations and procedures. Dev Cogn Neurosci. Aug 2018;32:16–22. doi:10.1016/j.dcn.2018.04.004

27. Karcher NR, Barch DM. The ABCD study: understanding the development of risk for mental and physical health outcomes. Neuropsychopharmacology. Jan 2021;46(1):131–142. doi:10.1038/s41386-020-0736-6

28. Compton WM, Dowling GJ, Garavan H. Ensuring the Best Use of Data: The Adolescent Brain Cognitive Development Study. JAMA Pediatr. Sep 1 2019;173(9):809–810. doi:10.1001/jamapediatrics.2019.2081

29. Rakesh D, Zalesky A, Whittle S. Assessment of Parent Income and Education, Neighborhood Disadvantage, and Child Brain Structure. JAMA Network Open. 2022;5(8):e2226208. doi:10.1001/jamanetworkopen.2022.26208

30. Van Dijk MT, Murphy E, Posner JE, Talati A, Weissman MM. Association of Multigenerational Family History of Depression With Lifetime Depressive and Other Psychiatric Disorders in Children. JAMA Psychiatry. 2021;78(7):778. doi:10.1001/jamapsychiatry.2021.0350

31. Singh GK. Area deprivation and widening inequalities in US mortality, 1969-1998. Am J Public Health. Jul 2003;93(7):1137–43. doi:10.2105/ajph.93.7.1137

32. Kind AJ, Jencks S, Brock J, et al. Neighborhood socioeconomic disadvantage and 30-day rehospitalization: a retrospective cohort study. Annals of internal medicine. 2014;161(11):765–774.

33. Acevedo-Garcia D, McArdle N, Hardy EF, et al. The child opportunity index: improving collaboration between community development and public health. Health Aff (Millwood). Nov 2014;33(11):1948–57. doi:10.1377/hlthaff.2014.0679

34. Investigation USDoJOoJPFBo. Data from: Uniform Crime Reporting Program Data: County-Level Detailed Arrest and Offense Data, United States, 2010. 2014. doi:10.3886/ICPSR33523.v2

35. Di Q, Amini H, Shi L, et al. Assessing NO(2) Concentration and Model Uncertainty with High Spatiotemporal Resolution across the Contiguous United States Using Ensemble Model Averaging. Environ Sci Technol. Feb 4 2020;54(3):1372–1384. doi:10.1021/acs.est.9b03358

36. Requia WJ, Di Q, Silvern R, et al. An Ensemble Learning Approach for Estimating High Spatiotemporal Resolution of Ground-Level Ozone in the Contiguous United States. Environ Sci Technol. Sep 15 2020;54(18):11037–11047. doi:10.1021/acs.est.0c01791

37. Fatemi F, Ardalan A, Aguirre B, Mansouri N, Mohammadfam I. Social vulnerability indicators in disasters: Findings from a systematic review. International journal of disaster risk reduction. 2017;22:219–227.

38. Ratcliffe M, Burd C, Holder K, Fields A. Defining rural at the US Census Bureau. American community survey and geography brief. 2016;1(8):1–8.

39. Ostermeijer F, Koster HRA, Van Ommeren J, Nielsen VM. Automobiles and urban density. Journal of Economic Geography. 2022;22(5):1073–1095. doi:10.1093/jeg/lbab047

40. Loewy RL, Pearson R, Vinogradov S, Bearden CE, Cannon TD. Psychosis risk screening with the Prodromal Questionnaire--brief version (PQ-B). Schizophr Res. Jun 2011;129(1):42–6. doi:10.1016/j.schres.2011.03.029

41. Dominguez MD, Wichers M, Lieb R, Wittchen HU, van Os J. Evidence that onset of clinical psychosis is an outcome of progressively more persistent subclinical psychotic experiences: an 8-year cohort study. Schizophrenia Bulletin. Jan 2011;37(1):84–93. doi:10.1093/schbul/sbp022

42. Palmer CE, Sheth C, Marshall AT, et al. A Comprehensive Overview of the Physical Health of the Adolescent Brain Cognitive Development Study Cohort at Baseline. Front Pediatr. 2021;9:734184. doi:10.3389/fped.2021.734184

43. Christensen GM, Li Z, Pearce J, et al. The complex relationship of air pollution and neighborhood socioeconomic status and their association with cognitive decline. Environ Int. Sep 2022;167:107416. doi:10.1016/j.envint.2022.107416

44. Carrico C, Gennings C, Wheeler DC, Factor-Litvak P. Characterization of Weighted Quantile Sum Regression for Highly Correlated Data in a Risk Analysis Setting. J Agric Biol Environ Stat. Mar 2015;20(1):100–120. doi:10.1007/s13253-014-0180-3

45. Czarnota J, Gennings C, Wheeler DC. Assessment of weighted quantile sum regression for modeling chemical mixtures and cancer risk. Cancer Inform. 2015;14(Suppl 2):159–71. doi:10.4137/cin.S17295

46. Renzetti S, Gennings C, Calza S. A weighted quantile sum regression with penalized weights and two indices. Frontiers in Public Health. 2023;11:1151821.

47. Bates D, Mächler M, Bolker B, Walker S. Fitting Linear Mixed-Effects Models Using lme4. Journal of Statistical Software. 10/07 2015;67(1):1–48. doi:10.18637/jss.v067.i01

48. Kuznetsova A, Brockhoff PB, Christensen RHB. lmerTest Package: Tests in Linear Mixed Effects Models. Journal of Statistical Software. 12/06 2017;82(13):1–26. doi:10.18637/jss.v082.i13

49. Tingley D, Yamamoto T, Hirose K, Keele L, Imai K. mediation: R Package for Causal Mediation Analysis. Journal of Statistical Software. 09/02 2014;59(5):1–38. doi:10.18637/jss.v059.i05

50. Renzetti S, Curtin P, Allan C, Bello G, Gennings C. gWQS: generalized weighted quantile sum regression. 2016;

51. Oh H, Nagendra A, Besecker M, Smith L, Koyanagi A, Wang JS. Economic strain, parental education and psychotic experiences among college students in the United States: Findings from the Healthy Minds Study 2020. Early Interv Psychiatry. Jul 2022;16(7):770–781. doi:10.1111/eip.13221

52. Koivukangas J, Tammelin T, Kaakinen M, et al. Physical activity and fitness in adolescents at risk for psychosis within the Northern Finland 1986 Birth Cohort. Schizophr Res. Feb 2010;116(2-3):152–8. doi:10.1016/j.schres.2009.10.022

53. Walker E, Mittal V, Tessner K. Stress and the hypothalamic pituitary adrenal axis in the developmental course of schizophrenia. Annu Rev Clin Psychol. 2008;4:189–216. doi:10.1146/annurev.clinpsy.4.022007.141248

54. Aberizk K, Collins MA, Addington J, et al. Life Event Stress and Reduced Cortical Thickness in Youth at Clinical High Risk for Psychosis and Healthy Control Subjects. Biol Psychiatry Cogn Neurosci Neuroimaging. Feb 2022;7(2):171–179. doi:10.1016/j.bpsc.2021.04.011

55. Aberizk K, Addington JM, Bearden CE, et al. Relations of Lifetime Perceived Stress and Basal Cortisol with Hippocampal Volume among Healthy Adolescents and those at Clinical High-Risk for Psychosis: A Structural Equation Modeling Approach. Biological Psychiatry. 2023/12// 2023:S0006322323017596. doi:10.1016/j.biopsych.2023.11.027

56. Aznar S, Jimenez-Zazo F, Romero-Blanco C, et al. Walkability and socio-economic status in relation to walking, playing and sports practice in a representative Spanish sample of youth: The PASOS study. PLoS One. 2024;19(3):e0296816. doi:10.1371/journal.pone.0296816

57. Glover TD, Todd J, Moyer L. Neighborhood Walking and Social Connectedness. Front Sports Act Living. 2022;4:825224. doi:10.3389/fspor.2022.825224

58. Zammit S, Lewis G, Rasbash J, Dalman C, Gustafsson J-E, Allebeck P. Individuals, Schools, and Neighborhood: A Multilevel Longitudinal Study of Variation in Incidence of Psychotic Disorders. Archives of General Psychiatry. 2010;67(9):914–922. doi:10.1001/archgenpsychiatry.2010.101

59. Bhavsar V, Boydell J, Murray R, Power P. Identifying aspects of neighbourhood deprivation associated with increased incidence of schizophrenia. Schizophrenia Research. 2014/06/01/ 2014;156(1):115–121. 10.1016/j.schres.2014.03.014

60. Newbury JB, Arseneault L, Beevers S, et al. Association of Air Pollution Exposure With Psychotic Experiences During Adolescence. JAMA Psychiatry. 2019;76(6):614–623. doi:10.1001/jamapsychiatry.2019.0056

61. St-Hilaire C, Brunila M, Wachsmuth D. High rises and housing stress: A spatial big data analysis of rental housing financialization. Journal of the American Planning Association. 2024;90(1):129–143.

62. Machlin L, Egger HL, Stein CR, et al. Distinct Associations of Deprivation and Threat With Alterations in Brain Structure in Early Childhood. Journal of the American Academy of Child and Adolescent Psychiatry. 2023;62(8):885–894.e3. doi:10.1016/j.jaac.2023.02.006

63. Thomas M, Rakesh D, Whittle S, Sheridan M, Upthegrove R, Cropley V. The neural, stress hormone and inflammatory correlates of childhood deprivation and threat in psychosis: A systematic review. Psychoneuroendocrinology. 2023/11/01/ 2023;157:106371. 10.1016/j.psyneuen.2023.106371

64. Vargas T, Rakhshan Rouhakhtar PJ, Schiffman J, Zou DS, Rydland KJ, Mittal VA. Neighborhood crime, socioeconomic status, and suspiciousness in adolescents and young adults at Clinical High Risk (CHR) for psychosis. Schizophr Res. Jan 2020;215:74–80. doi:10.1016/j.schres.2019.11.024

65. Wilson C, Smith ME, Thompson E, et al. Context matters: The impact of neighborhood crime and paranoid symptoms on psychosis risk assessment. Schizophr Res. Mar 2016;171(1-3):56–61. doi:10.1016/j.schres.2016.01.007

66. Zewdie H, Zhao AY, Patel HH, et al. The association between neighborhood quality, youth physical fitness, and modifiable cardiovascular disease risk factors. Ann Epidemiol. May 2021;57:30–39. doi:10.1016/j.annepidem.2021.02.004

67. Chaix B, Kestens Y, Perchoux C, Karusisi N, Merlo J, Labadi K. An Interactive Mapping Tool to Assess Individual Mobility Patterns in Neighborhood Studies. American Journal of Preventive Medicine. 2012/10/01/ 2012;43(4):440–450. 10.1016/j.amepre.2012.06.026

68. Mittal VA, Vargas T, Osborne KJ, et al. Exercise Treatments for Psychosis: A Review. Curr Treat Options Psychiatry. Jun 2017;4(2):152–166. doi:10.1007/s40501-017-0112-2

69. Fish JS, Ettner S, Ang A, Brown AF. Association of perceived neighborhood safety with [corrected] body mass index. Am J Public Health. Nov 2010;100(11):2296–303. doi:10.2105/ajph.2009.183293

70. Leyden KM, Hogan MJ, D’Arcy L, Bunting B, Bierema S. Walkable Neighborhoods. Journal of the American Planning Association. 2024/01/02 2024;90(1):101-114. doi:10.1080/01944363.2022.2123382

## References

1. Tingley D, Yamamoto T, Hirose K, Keele L, Imai K. mediation: R Package for Causal Mediation Analysis. Journal of Statistical Software. 09/02 2014;59(5):1–38. doi:10.18637/jss.v059.i05

