## Supplementary Materials for "Exposure profiles of social-environmental neighborhood factors and psychotic-like experiences"

**Supplemental Online Content**

**eMethods.**

**eTable 1**. Sociodemographic characteristics of included and excluded participants.

**eTable 2**. List of exposure definitions and years measured.

**eTable 3**. Categorization of team and individual sports.

**eTable 4**. Sensitivity subgroup analysis of associations between exposure profiles and persistent distressing PLE.

**eTable 5.** Variance inflation factors for the exposure profiles and covariates.

**eTable 6**. Subgroup mediation analysis of physical activity in the association between exposure profiles and persistent distressing PLE.

**eTable 7**. Subgroup mediation analysis of individual sports in the association between exposure profiles and persistently distressing PLE.

**eTable 8**. Mean weights of individual exposure components for positively constrained weighted quantile sum regression.

**eTable 9**. Mean weights of individual exposure components for negatively constrained weighted quantile sum regression.

**eFigure 1**. Flowchart of missing values.

**eFigure 2**. Bivariate correlation matrix for all exposure components.

**eReferences.**

**eMethods.**

**Mediation analyses**

For the mediation analysis, total physical activity and subcategories of physical activity (i.e., team and individual sports) were tested as mediators for the relationship between only the subgroups of significant exposure profiles and persistent distressing PLE. Linear mixed models tested the association between the exposure profile (in comparison to the reference profile (Profile 1) and mediators, and logistic mixed models assessed the association between mediators and persistent distressing PLE. Direct effects were determined using a logistic mixed regression model adjusted for mediators and covariates, while total effects used the original model adjusted for covariates. The indirect effect (i.e., average causal mediation effect) is then estimated with 5000 bootstraps using the R package “mediation.” This general procedure is based on Monte Carlo simulation, which when computing said indirect effect, accommodates for differences in the distributions of both the mediator and the outcome.^1,2^

**eTable 1. Sociodemographic characteristics of included and excluded participants.**

A. **Study sample characteristics. B. Area-level Deprivation Index. C. Child Opportunity Index. D. Crime. E. Environmental Quality. F. Social Vulnerability Index. G. Urbanicity Features. H. Physical Activities. I. Outcome Variable.**

|  | Total: 11868 | |  |
| --- | --- | --- | --- |
| Variables | Included | Excluded | P ^a^ |
| **A. Sample Characteristics** | | |  |
| N (%) | 8,145 (68.6%) | 3,723 (31.4%) |  |
| Age (mean (SD)) | 9.92 (0.63) | 9.91 (0.62) | 0.79 |
| Female Sex (%) ^b^ | 3,868 (47.5) | 339 (47.2) | < .001 |
| Race/Ethnicity (%) |  |  | < .001 |
| Non-Hispanic White | 4,711 (57.8) | 1,462 (39.3) |  |
| Non-Hispanic Black | 912 (11.2) | 872 (23.4) |  |
| Non-Hispanic Asian | 169 ( 2.1) | 83 ( 2.2) |  |
| Non-Hispanic Others ^c^ | 873 (10.7) | 374 (10.1) |  |
| Hispanic Ethnicity | 1,480 (18.2) | 930 (25.0) |  |
| Parent Bachelor's Degree (%) | 5,401 (66.3) | 263 (37.1) | < .001 |
| Income-to-needs ratio (mean (SD)) | 4.34 (3.31) | 3.59 (3.32) | < .001 |
| Family History of Psychosis (%) | 668 (8.2) | 49 ( 6.8) | 0.22 |
| **B. Area-level Deprivation Index - Median (IQR)** | | |  |
| % Single Parent Households | 13.51 [8.62, 21.46] | 20.49 [11.94, 29.95] | < .001 |
| % Home Ownership | 71.91 [54.68, 83.94] | 58.18 [38.66, 77.90] | < .001 |
| % Less Than 9 Years Education | 2.19 [0.92, 5.06] | 4.32 [1.85, 10.83] | < .001 |
| % At least High School Education | 92.89 [86.29, 96.40] | 87.25 [74.69, 93.94] | < .001 |
| % White Collar Occupation | 94.50 [91.68, 96.63] | 94.24 [90.89, 96.54] | < .001 |
| Median Family Income | 75,065.00 [54,375.00, 98,690.00] | 56,047.00 [38,070.25, 79,467.25] | < .001 |
| Income Disparity | 1.96 [1.23, 2.79] | 2.57 [1.58, 3.53] | < .001 |
| Median Property Value | 227,200.00 [154,200.00, 321,800.00] | 203,600.00 [116,550.00, 296,450.00] | < .001 |
| Median Gross Rent | 1,056.00 [857.00, 1,332.00] | 1,017.50 [851.50, 1,245.25] | 0.004 |
| Median Monthly Mortgage | 1,403.00 [1,083.00, 1,723.00] | 1,299.50 [945.00, 1,648.75] | < .001 |
| Crowding | 1.53 [0.48, 3.49] | 2.80 [0.71, 6.27] | < .001 |
| Unemployment | 7.22 [4.89, 10.46] | 9.61 [6.34, 14.22] | < .001 |
| % Below Poverty Line | 6.53 [3.07, 13.42] | 12.71 [5.11, 24.52] | < .001 |
| % Below 138% Poverty Line | 15.01 [8.90, 26.63] | 25.21 [12.67, 40.27] | < .001 |
| % Households with No Car | 4.60 [2.10, 9.64] | 7.49 [3.49, 14.54] | < .001 |
| % Poor Plumbing | 0.00 [0.00, 0.29] | 0.00 [0.00, 0.50] | < .001 |
| **C. Child Opportunity Index - Median (IQR)** | | |  |
| Industrial Pollutants | 0.40 [-0.45, 0.71] | 0.45 [-0.57, 0.74] | 0.40 |
| Hazardous Waste Dump Sites | 0.27 [0.27, 0.27] | 0.27 [0.27, 0.27] | 0.06 |
| Access to Food | 0.38 [-0.16, 0.67] | 0.32 [-0.54, 0.67] | 0.04 |
| Access to Green Space | -0.30 [-0.90, 0.55] | -0.63 [-1.35, 0.13] | < .001 |
| Walkability | 10.50 [7.17, 14.00] | 12.00 [7.83, 14.67] | < .001 |
| **D. Crime - Median (IQR)** | | |  |
| Total Crime | 22,761.33 [7,832.00, 53,399.67] | 31,626.67 [10,664.33, 77,667.00] | < .001 |
| **E. Environmental Quality -Median (IQR)** | | |  |
| PM2.5 (μg/m3) | 7.68 [6.56, 8.60] | 8.19 [6.93, 9.02] | < .001 |
| NO2 (ppb) | 18.74 [14.75, 22.19] | 19.01 [15.50, 22.30] | 0.17 |
| O3 (ppb) | 40.43 [38.17, 45.13] | 40.65 [38.40, 45.95] | 0.06 |
| Lead Risk | 15.73 [6.82, 31.35] | 20.63 [8.41, 37.54] | < .001 |
| Proximity to Roadways | 855.45 [390.82, 1,575.26] | 766.20 [404.30, 1,506.20] | 0.27 |
| **F. Social Vulnerability Index - Median (IQR)** | | |  |
| % Ethnoracial Minority | 46.48 [28.45, 70.43] | 70.34 [45.12, 89.35] | < .001 |
| % Non-English Speakers | 48.57 [27.67, 71.35] | 64.75 [33.95, 89.34] | < .001 |
| **G. Urbanicity Features - Median (IQR)** | |  |  |
| Population Density | 1,627.60 [786.47, 2,670.60] | 1,795.09 [853.96, 3,253.67] | < .001 |

**H. Potential Mediators - Median (IQR)**

| Total Physical Activities | 2.37 [0.87, 4.50] | 1.69 [0.00, 4.07] | < .001 |
| --- | --- | --- | --- |
| Team Sports | 0.75 [0.00, 2.17] | 0.33 [0.00, 1.83] | < .001 |
| Individual Sports | 0.92 [0.00, 2.29] | 0.37 [0.00, 2.00] | < .001 |
| **I. Outcome Variable** | | |  |
| Persistent Distressing PLE (%) | 1,605 (19.7) | 419 (23.7) | < .001 |

^a^ X^2^ tests were used for categorical variables and Kruskal–Wallis tests were used for non-normal continuous variables.

^b^ sex=3 (n=3,0.4%) is excluded.

^c^ Others included participants who reported a race that was not included in the list, did not know their race, or did not disclose.

**eTable 2. List of exposure definitions and years measured. ^a^**

| **Index Domain**/Exposure Component | **Definition** | **Year(s) Measured** |
| --- | --- | --- |
| **Area-level Deprivation Index** | |  |
| % Single Parent Households | Percentage of single-parent households | 2010-2014 |
| % Home Ownership | Percentage of owner-occupied households | 2010-2014 |
| % Less Than 9 Years Education | Percentage of population aged >=25 years with <9 years of education | 2010-2014 |
| % At least High School Education | Percentage of population aged >=25 years with at least a high school diploma | 2010-2014 |
| % White Collar Occupation | Percentage of employed persons aged >=16 years in white-collar occupations | 2010-2014 |
| Median Family Income | Median family income | 2010-2014 |
| Income Disparity | Income disparity is defined by Singh (2003) as the log of 100 x ratio of the number of households with <10000 annual income to the number of households with >50000 annual income | 2010-2014 |
| Median Home Value | Median home value | 2010-2014 |
| Median Gross Rent | Median gross rent | 2010-2014 |
| Median Monthly Mortgage | Median monthly mortgage | 2010-2014 |
| Crowding | Percentage of occupied housing units with >1 person per room (crowding) | 2010-2014 |
| Unemployment | Percentage of civilian labor force population aged >=16 y unemployed (unemployment rate) | 2010-2014 |
| % Below Poverty Line | Percentage of families below the poverty level | 2010-2014 |
| % Below 138% Poverty Line | Percentage of population below 138% of the poverty threshold | 2010-2014 |
| % Households with No Car | Percentage of occupied housing units without a motor vehicle | 2010-2014 |
| % Poor Plumbing | Percentage of occupied housing units without complete plumbing (log) | 2010-2014 |
| **Child Opportunity Index 2.0** | |  |
| Industrial Pollutants | Index of toxic chemicals released by industrial facilities, converted to natural log units, transformed to z-scores and multiplied by -1 | 2015 |
| Hazardous Waste Sites | Average number of Superfund sites within a 2-mile radius, converted to natural log units, transformed to z-scores and multiplied by -1 | 2015 |
| Access to Food | Percentage households without a car located further than a half-mile from the nearest supermarket, transformed to z-scores and multiplied by -1 | 2015 |
| Access to Green Space | Percentage impenetrable surface areas such as rooftops, roads or parking lots, transformed to z-scores and multiplied by -1 | 2015 |
| Walkability | National Walkability Index from the Smart Location Database created by the United States Environmental Protection Agency (https://www.epa.gov/smartgrowth/smart-location-mapping#walkability) | 2010 |
| **Crime** |  |  |
| Total Crime | County-level counts of arrests and offenses from Uniform Crime Reporting Program Data (https://doi.org/10.3886/ICPSR33523.v2). | average from 2010 -2012 |
| **Environmental Quality** |  |  |
| PM2.5 (μg/m3) | Spatio-temporal model predictions measured in μg/m3 at 1 km2 resolution | 2016 |
| NO2 (ppb) | Spatio-temporal model predictions measured in ppb (parts per billion) at 1 km2 resolution | 2016 |
| O3 (ppb) | Spatio-temporal model predictions measured in ppb (parts per billion) at 1 km2 resolution | 2016 |
| Lead Risk | Estimated percentage of homes at risk for lead exposure given lead-based paint in census tract of primary residential address | 2010-2014 |
| Proximity to Roadways | Number of meters away from major road or highway | 2016 |
| **Social Vulnerability Index** | |  |
| Percent Ethnoracial Minority | Percentage of ethnoracial minority population (i.e., all but white, non-Hispanic) | 2014-2018 |
| Percent Non-English Speakers | Percentage of persons at least 5 years old who speak English “less than well” | 2014-2018 |
| **Urbanicity Features** |  |  |
| Population Density | Census tract population count adjusted to match the 2015 Revision of UN WPP Country Totals in persons per 1 km2. | 2010 |

^a^ A total of 29 exposure variables from the Area-level Deprivation Index, Child Opportunity Index 2.0, crime, environmental quality, and social vulnerability index were used to create exposure profiles.

**eTable 3. Categorization of team and individual sports.^a^**

| **Team Sports** | **Individual Sports** |
| --- | --- |
| Softball, Baseball | Dance, ballet |
| Basketball | Climbing |
| Field Hockey | Gymnastics |
| Football | Horse-riding, polo |
| Ice Hockey | Ice or Inline Skating |
| Lacrosse | Martial Arts |
| Rugby | Skateboarding |
| Soccer | Snowboarding, Skiing |
| Volleyball | Surfing  Swimming |
|  | Tennis |
|  | Track, running, cross-country |
|  | Wrestling |
|  | Yoga, Tai Chi |

^a^ A data-driven approach to characterize team and individual sports is further described in a prior report and has been used in prior literature.^3,4^

**eTable 4. Sensitivity subgroup analysis of associations between exposure profiles and persistent distressing PLE.**

|  |  | **Model A (N = 8145) ^a^** | | | **Model B (N = 7143) ^b^** | | |
| --- | --- | --- | --- | --- | --- | --- | --- |
|  | **Features/Descriptions** | **OR** | **95% CI** | ***P*** | **OR** | **95% CI** | ***P*** |
| **Exposure** |  |  |  |  |  |  |  |
| Profile 1 | Suburban affluent areas (reference) | | | |  |  |  |
| Profile 2 | Suburban areas with high pollutants | 1.04 | 0.89—1.22 | 0.62 | 1.10 | 0.92—1.31 | 0.29 |
| Profile 3 | Rural areas with low walkability and high ozone | 1.25 | 1.03—1.53 | 0.02 | 1.27 | 1.02—1.60 | 0.03 |
| Profile 4 | Urban areas with high ADI, high crime, and high pollution | 1.29 | 1.01—1.65 | 0.04 | 1.32 | 1.00—1.74 | 0.05 |
| Profile 5 | Urban areas with high ADI and low access to food | 1.30 | 1.03—1.66 | 0.03 | 1.23 | 0.93—1.62 | 0.14 |

**^a^** Model A: sensitivity analysis for another version of PLE where participants who met distressing criteria at the last follow-up visit were included in the persistently distressing PLE group.

**^b^** Model B: sensitivity analysis for a subgroup of participants who indicated they have lived in their current address for more than 1 year.

Note: All models treated site and family as two random intercepts and adjusted for individual-level covariates: age, sex, race/ethnicity, family history of psychosis, parents with bachelor’s degrees, and income-to-needs ratio.

**eTable 5. Variance inflation factors for exposure profiles and covariates.**

|  | **Variance Inflation Factor** |
| --- | --- |
| Exposure profile | 1.048572 |
| Age | 1.001157 |
| Sex | 1.001355 |
| Family history of psychosis | 1.004164 |
| Race/ethnicity | 1.041209 |
| Parental college education | 1.174771 |
| Income-to-needs ratio | 1.202346 |

**eTable 6. Subgroup mediation analysis of physical activity in the association between significant exposure profiles and persistent distressing PLE. ^a^**

| **Variable/Effect** | **Beta** | **95% CI** | **P** | **Proportion Mediated** |
| --- | --- | --- | --- | --- |
| **Profile 3 compared to Profile 1 (n = 3980)** |  |  |  |  |
| **Direct Effect** |  |  |  |  |
| Profile 3 -> Persistent distressing PLE | 0.028 | <-0.001—0.060 | 0.052 |  |
| **Total Effect** |  |  |  |  |
| Profile 3 -> Persistent distressing PLE | 0.030 | 0.001—0.060 | 0.040 |  |
| **Individual Paths** |  |  |  |  |
| Profile 3-> Physical Activity | -0.138 | -0.216—-0.060 | 0.001 |  |
| Physical Activity -> Persistent distressing PLE | -0.089 | -0.182—0.004 | 0.060 |  |
| **Indirect Effect** |  |  |  |  |
| Profile 3 -> Physical Activity -> Persistent PLE | 0.002 | < -.001—0.002 | 0.071 | 4.93% |
| **Profile 4 compared to Profile 1 (n = 3236)** |  |  |  |  |
| **Direct Effect** |  |  |  |  |
| Profile 4 -> Persistent distressing PLE | 0.038 | -0.003—0.080 | 0.069 |  |
| **Total Effect** |  |  |  |  |
| Profile 4 -> Persistent distressing PLE | 0.039 | -0.002—0.080 | 0.064 |  |
| **Individual Paths** |  |  |  |  |
| Profile 4 -> Physical Activity | -0.172 | -0.287—-0.057 | 0.003 |  |
| Physical Activity -> Persistent distressing PLE | -0.043 | -0.143—0.057 | 0.399 |  |
| **Indirect Effect** |  |  |  |  |
| Profile 4 -> Physical Activity -> Persistent PLE | 0.001 | -0.002—0.004 | 0.478 | 1.94% |

^a^ Mediation analysis included recruiting sites as a random intercept and adjusted individual covariates: age, sex, race, family history of psychosis, parents with a bachelor’s degree, and income-to-needs ratio. Physical activity was not significantly associated with persistently distressing PLE in these two subsamples. Total physical activities did not mediate the association between Profile 3 or Profile 4 and persistent distressing PLE.

**eTable 7. Subgroup mediation analysis of individual sports in the association between exposure profiles and persistent distressing PLE. ^a^**

| **Variable/Effect** | **Beta** | **95% CI** | **P** | **Proportion Mediated** |
| --- | --- | --- | --- | --- |
| **Profile 3 compared to Profile 1 (n = 3980)** |  |  |  |  |
| **Direct Effect** |  |  |  |  |
| Profile 3 -> Persistently distressing PLE | 0.030 | 0.002—0.060 | 0.032 |  |
| **Total Effect** |  |  |  |  |
| Profile 3 -> Persistent PLE | 0.030 | 0.002—0.059 | 0.033 |  |
| **Individual Paths** |  |  |  |  |
| Profile 3-> Individual Sports | -0.073 | -0.154—0.008 | 0.078 |  |
| Individual Sports -> Persistent PLE | 0.021 | -0.062—0.103 | 0.625 |  |
| **Indirect Effect** |  |  |  |  |
| Profile 3 -> Individual Sports -> Persistent PLE | < .001 | -0.001—0.002 | 0.915 | 0.17% |
| **Profile 4 compared to Profile 1 (n = 3236)** |  |  |  |  |
| **Direct Effect** |  |  |  |  |
| Profile 4 -> Persistently distressing PLE | 0.038 | -0.003—0.080 | 0.070 |  |
| **Total Effect** |  |  |  |  |
| Profile 4 -> Persistently distressing PLE | 0.039 | -0002—0.079 | 0.064 |  |
| **Individual Paths** |  |  |  |  |
| Profile 4 -> Individual Sports | -0.136 | -0.253—-0.019 | 0.023 |  |
| Individual Sports -> Persistently distressing PLE | 0.032 | -0.060—0.125 | 0.493 |  |
| **Indirect Effect** |  |  |  |  |
| Profile 4 -> Individual Sports -> Persistent PLE | < .001 | -0.001—0.001 | 0.481 | 1.93% |

^a^ Mediation analysis included recruiting sites as a random intercept and adjusted individual covariates: age, sex, race, family history of psychosis, parents with a bachelor’s degree, and income-to-needs ratio. Individual sports were not significantly associated with persistently distressing PLE in these two subsamples. Individual sports did not mediate the association between Profile 3 or Profile 4 and persistent distressing PLE.

**eTable 8. Mean weights of individual exposure components for positively constrained weighted quantile sum regression.^a^**

| **Abbreviation** | **Full Name** | **Mean Weight** |
| --- | --- | --- |
| TotalCrime | Total Crime | 0.130 |
| Walkability | Walkability | 0.106 |
| MinorityConcentration | % Ethnoracial Minority | 0.090 |
| O3 | O3 | 0.085 |
| IndustrialPollutants | Industrial Pollutants | 0.077 |
| Income | Median Family Income | 0.075 |
| WhiteCollar | % White Collar | 0.057 |
| Crowding | Crowding | 0.050 |
| Unemployment | Unemployment | 0.050 |
| NonEnglishSpeakers | % Non-English Speakers | 0.045 |
| SuperfundSites | Hazardous Waste Dump Sites | 0.044 |
| LessThan9YrsEd | % Less Than 9 Years Education | 0.037 |
| NO2 | NO2 | 0.024 |
| HomeOwnership | % Home Ownership | 0.021 |
| PM25 | PM2.5 | 0.020 |
| Proxtoroad | Proximity to Roadways | 0.018 |
| PoorPlumbing | % Poor Plumbing | 0.012 |
| LeadRisk | Lead Risk | 0.011 |
| HighSchoolEd | % High School Education | 0.010 |
| Mortgage | Median Monthly Mortgage | 0.009 |
| SingleParent | % Single-Parent Household | 0.009 |
| Rent | Median Gross Rent | 0.007 |
| BelowPoverty | Below Poverty Line | 0.006 |
| PropertyValue | Median Property Value | 0.004 |
| GreenSpace | Access to Green Space | 0.002 |
| Below138Poverty | % Below 138% Poverty Line | 0.002 |
| FoodAccess | Access to Food | 0.001 |
| NoCar | % Household with no Car | < .001 |
| IncomeDisparity | Income Disparity | < .001 |

^a^ The positive constrained WQS model for the exposure mixture and persistent distressing PLE is significant (adjusted OR: 1.12, 95% CI: 1.02—1.23, p = 0.03). The positive constrained weighted quantile sum adjusted for individual-level covariates: age, sex, race/ethnicity, family history of psychosis, parents with bachelor’s degrees, income-to-needs ratio, and recruiting sites. The results presented have been bootstrapped at 1000.

**eTable 9. Mean weights of individual exposure components for negatively constrained weighted quantile sum regression.^a^**

| **Abbreviation** | **Full Name** | **Mean Weight** |
| --- | --- | --- |
| PoorPlumbing | % Poor Plumbing | 0.318 |
| FoodAccess | Access to Food | 0.190 |
| Walkability | Walkability | 0.162 |
| PM25 | PM2.5 | 0.095 |
| Proxtoroad | Proximity to Roadways | 0.067 |
| GreenSpace | Access to Green Space | 0.029 |
| NO2 | NO2 | 0.023 |
| IncomeDisparity | Income Disparity | 0.022 |
| NoCar | % Household with no Car | 0.021 |
| SuperfundSites | Hazardous Waste Dump Sites | 0.020 |
| O3 | O3 | 0.019 |
| LeadRisk | Lead Risk | 0.013 |
| WhiteCollar | % White Collar | 0.007 |
| IndustrialPollutants | Industrial Pollutants | 0.005 |
| Rent | Median Gross Rent | 0.004 |
| TotalCrime | Total Crime | 0.002 |
| Mortgage | Median Monthly Mortgage | 0.001 |
| PropertyValue | Median Property Value | 0.001 |
| NonEnglishSpeakers | % Non-English Speakers | < .001 |
| Crowding | Crowding | < .001 |
| SingleParent | % Single-Parent Household | < .001 |
| LessThan9YrsEd | % Less Than 9 Years Education | < .001 |
| HighSchoolEd | % High School Education | < .001 |
| HomeOwnership | % Home Ownership | < .001 |
| BelowPoverty | Below Poverty Line | < .001 |
| Below138Poverty | % Below 138% Poverty Line | < .001 |
| Unemployment | Unemployment | < .001 |
| Income | Median Family Income | < .001 |
| MinorityConcentration | % Ethnoracial Minority | < .001 |

^a^ The negative constrained WQS model for the exposure mixture and persistent distressing PLE is not significant (adjusted OR: 0.99, 95%: 0.90—1.09, p = 0.95). The negative constrained weighted quantile sum adjusted for individual-level covariates: age, sex, race/ethnicity, family history of psychosis, parents with bachelor’s degrees, income-to-needs ratio, and recruiting sites. The results presented have been bootstrapped at 1000.

**eFigure 1. Flowchart of missing values.^a^**

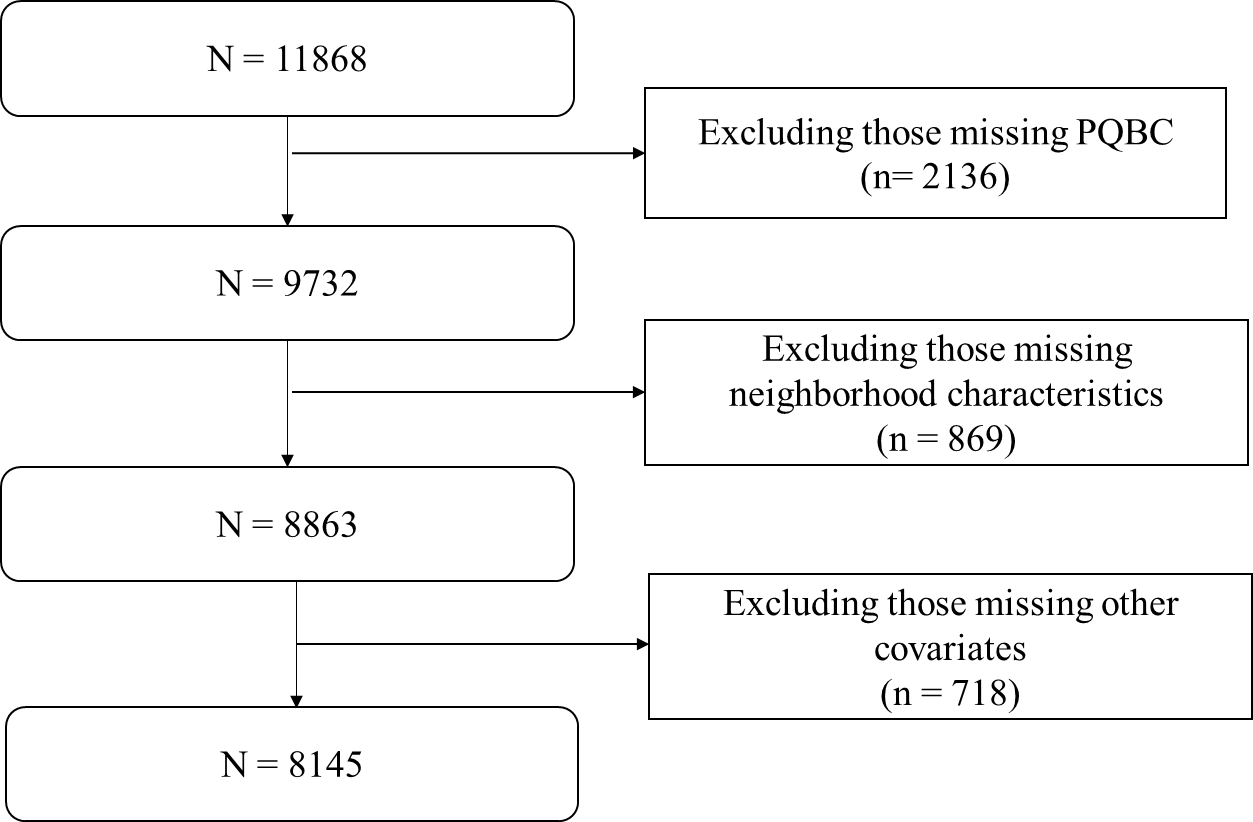

Abbreviations: PQBC, Prodromal Questionnaire-Brief Child Version.

^a^ Other covariates include exclusion of sex = 3 (n = 3), missing parental education (n = 9), and missing household income to calculate income-to-needs ratio (n = 706).

**eFigure 2. Bivariate correlation matrix for all exposure components.^a^**
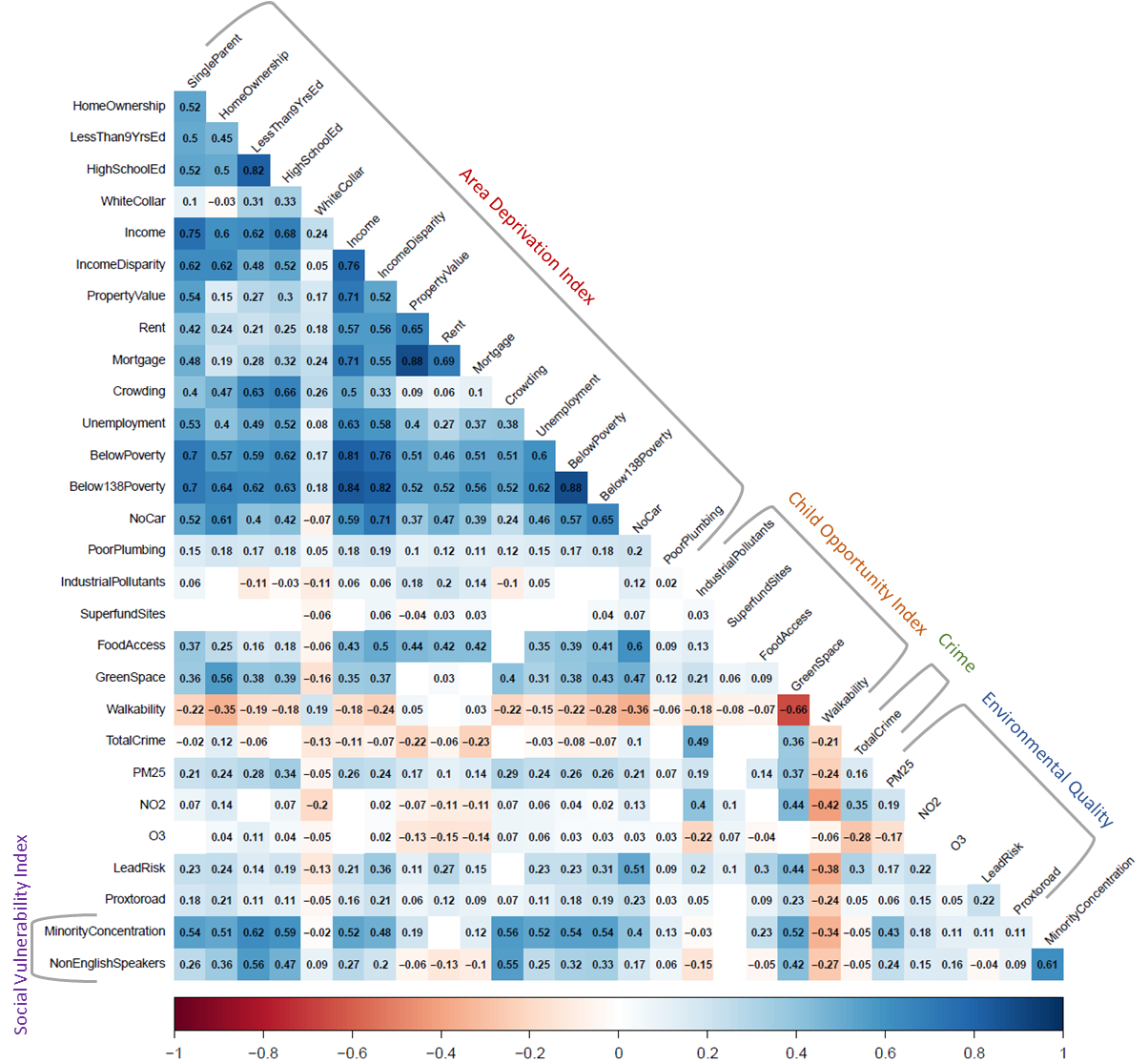

^a^ Bivariate correlation matrix for all exposure components, organized into five domains indicated by gray brackets and labeled with colored texts. All variables were coded such that higher scores indicate worse outcomes. Each cell displays the Pearson correlation coefficient for significant correlation between variables (p < 0.05).

**eReferences**

1.	Tingley D, Yamamoto T, Hirose K, Keele L, Imai K. mediation: R Package for Causal Mediation Analysis. *Journal of Statistical Software*. 09/02 2014;59(5):1 - 38. doi:10.18637/jss.v059.i05

2.	Tingley D, Yamamoto T, Hirose K, Keele L, Imai K, Yamamoto MT. Package ‘mediation’. *Computer software manual*. 2019:175-184.

3.	Conley MI, Hindley I, Baskin-Sommers A, Gee DG, Casey BJ, Rosenberg MD. The importance of social factors in the association between physical activity and depression in children. *Child Adolesc Psychiatry Ment Health*. 2020;14:28. doi:10.1186/s13034-020-00335-5

4.	Kunitoki K, Hughes D, Elyounssi S, et al. Youth Team Sports Participation Associates With Reduced Dimensional Psychopathology Through Interaction With Biological Risk Factors. *Biological Psychiatry Global Open Science*. 2023/02/10/ 2023;doi:<https://doi.org/10.1016/j.bpsgos.2023.02.001>
